# The tumor suppressor Adenomatous polyposis coli regulates T lymphocyte migration. Insights from familial polyposis patients

**DOI:** 10.1101/2021.06.21.21259262

**Authors:** Marta Mastrogiovanni, Pablo Vargas, Thierry Rose, Céline Cuche, Marie Juzans, Elric Esposito, Hélène Laude, Charlotte Renaudat, Marie-Noëlle Ungeheuer, Jérôme Delon, Andrés Alcover, Vincenzo Di Bartolo

**Author notes:** Correspondence: Andrés Alcover, Department of Immunology, Institut Pasteur, 25-28 rue Docteur Roux, 75015 Paris; and Vincenzo Di Bartolo, Department of Immunology, Institut Pasteur, 25-28 rue Docteur Roux, 75015 Paris. The authors have declared that no conflict of interest exists.

## Abstract

Adenomatous polyposis coli (APC) is a tumor suppressor whose mutations underlie familial adenomatous polyposis (FAP) and colorectal cancer. Although its role in intestinal epithelial cells is well characterized, APC importance for anti-tumor immunity is ill defined. APC regulates cytoskeleton organization, cell polarity and migration in various cells types. Here we address whether APC plays a role in T lymphocyte migration, a key step of anti-tumor immune responses.

Using a series of cell biology tools, we demonstrated that T cells from FAP patients carrying APC mutations display adhesion and migration defects. Concomitantly, they presented lower expression of the integrin VLA-4. To further dissect the cellular mechanisms underpinning these defects, we depleted APC in the CEM T cell line. We found that APC is critical not only for VLA-4-dependent adhesion but also for actomyosin and microtubule organization in migrating T cells. Finally, APC-silenced CEM cells preferentially adopt an ameboid-like migration featuring unstructured pseudopodia and blebbing.

These findings underscore a role of APC in T cell migration *via* modulation of integrin-dependent adhesion and cytoskeleton reorganization. Hence, APC mutations in FAP patients not only drive intestinal neoplasms, but also impair T cell migration, potentially leading to inefficient T cell-mediated anti-tumor immunity.

## Introduction

Familial adenomatous polyposis (FAP) is an autosomal dominant inherited disease that often results from germline mutations in the adenomatous polyposis coli (APC) gene, a tumor suppressor and polarity regulator. APC protein regulates the Wnt/β-catenin signaling pathway as a component of the β-catenin destruction complex. This pathway is crucial for intestinal epithelium homeostasis. APC mutations alter epithelial cell differentiation, proliferation, polarization, and migration, thus disrupting tissue architecture (1). FAP patients carrying APC gene mutations develop numerous pre-cancerous colorectal polyps growing from the age of 10–12 years, with high risk of cancer by the age of 40 (2). Thus, APC alterations are prerequisite for progression towards malignancy.

APC is a polarity regulator also necessary for polarized cell migration (3). Through its N-terminal and C-terminal domains, APC interacts with both actin and microtubule regulatory molecules controlling microtubule network organization and dynamics and actin polymerization (4–6). Finally, APC controls the organization of intermediate filaments (7).

The key role of immunity in human colorectal carcinoma was underscored by the correlation between the type and frequency of infiltrating T lymphocytes and favorable patient prognosis (8, 9). Once activated in lymph nodes, lymphocytes go through bloodstream recirculation, adhesion-dependent trans-endothelial migration to finally invade tumor tissues, where they execute their effector functions to eliminate tumor cells. Lymphocyte migration through endothelium and tissues occurs in response to chemokine cues and relies on the coordination of acto-myosin and microtubule cytoskeleton dynamics, and of integrin-mediated adhesion (6, 10, 11). T cell migration involves the formation of a lamellipodium at the leading edge and a uropod at the trailing edge. Integrins, as LFA-1 (α_L_β_2_) and VLA-4 (α_4_β_1_), concentrate at various cellular areas, ensuring firm adhesion by binding to ICAM-1 and VCAM-1 on vascular endothelial cells and to fibronectin in extracellular matrix. Various adhesion molecules (i.e. CD44, ICAM-1,2,3, etc.) concentrate at the uropod supporting cell-cell interactions. Integrins and uropod adhesion molecules are linked to the cortical actin cytoskeleton *via* talin and ezrin and moesin proteins, respectively (12–14). Cell polarity complexes orchestrate the cytoskeletal crosstalk needed for directional migration (6).

Our recent work highlighted the association of APC with microtubules in T cells (15, 16) and showed that APC defects impair cytoskeleton organization at immunological synapses of CD4 and CD8 T cells, compromising differentiation and anti-inflammatory function of intestinal T regulatory cells (15), and cytotoxic activity of CD8 T cells (16). This revealed new roles of APC in T cell effector functions crucial for anti-tumor immunity.

Here, we unveiled that T lymphocytes from FAP patients carrying APC mutations show defective chemokine-induced migration through micropores or human umbilical vein endothelial cell (HUVEC) monolayers. Defects of FAP patients’ cells were maintained when migrating through chemokine-free, fibronectin-coated microchannels, indicating a switch from lamellipodium-to bleb-driven migration. Moreover, VLA-4 integrin expression was diminished in FAP patients T cells likely contributing to their defective adhesion to VCAM-1- or fibronectin-coated surfaces. Finally, studying an APC-silenced T cell line as a complementary model of APC defects, we revealed that APC-silenced cells displayed less structured lamellipodia and impaired filopodia formation. Altogether, we show a concomitant impact of APC defects on adhesion and cytoskeleton organization resulting in impaired T cell migration.

Therefore, APC mutations in FAP patients may weaken both intestinal epithelium homeostasis and anti-tumor immune surveillance, that relies on the ability of the immune cells to migrate to and through transformed tissues.

## Results

### Adhesion- and friction-dependent migration is impaired in T lymphocytes from patients carrying APC mutations

In non-lymphoid cells, as astrocytes, APC is at the crossroads of cytoskeleton and mechanical forces necessary to promote polarized cell migration (3). We hypothesized that APC is involved in T lymphocyte migration that may be impaired in cells from FAP patients carrying APC mutations. Therefore, we assessed the ability of 5-days activated T cells from FAP patients *versus* age and sex-matched healthy donors to migrate through transwell filters in response to CXCL12 chemokine. Chemokine-induced migration of both CD4 and CD8 T cells from FAP patients was significantly impaired as compared to their matched controls (Figure 1A).

**Figure 1:**
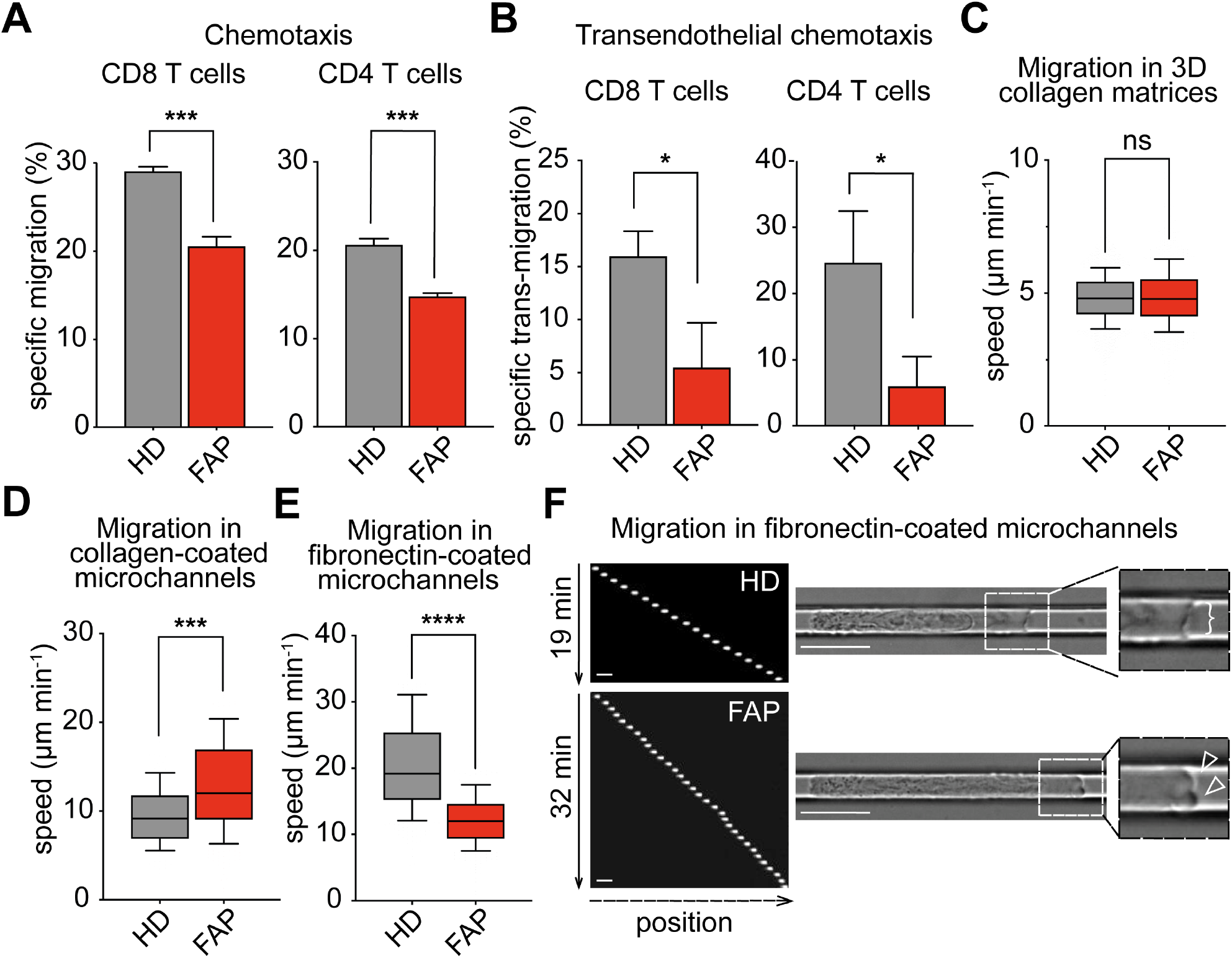
T cells from FAP patients present migration defects. **(A)** Transmigration through transwell filters was assessed for T cells from FAP patients (FAP) and matched healthy donors (HD). Bars represent mean + SEM (6 individual pairs: FAP01 to FAP06) of specific migration of CD8 and CD4 T cells. Statistical differences were calculated by paired t-test. *** = p<0.001. **(B-F)** T cells from FAP patients and matched healthy donors were re-activated after thawing and their transwell migration through HUVEC monolayers was assessed **(B)**. Bar graphs represent mean + SEM (3 individual pairs: FAP01, FAP02, FAP07) of specific migration of CD8 and CD4 T cells. Statistical differences were calculated by paired t-test. * = p<0.05. **(C-F)** CD8 T cells were sorted after thawing and were assessed for their migration in absence of chemokine stimuli in 3D collagen gels **(C)** or in microchannels **(D-F)**. Boxes illustrate 10-90 percentiles of values, and whiskers represent the range of values. Statistical differences were calculated by Mann-Whitney unpaired test.*** p<0.001, **** p<0.0001, ns: not significant. **(C)** CD8 T cell migration in 3D collagen matrices was recorded by time-lapse video microscopy. Mean speed of migrating cells from 4 individual pairs (FAP01, FAP03, FAP05, FAP07) is shown. **(D)** CD8 T cell migration through collagen type I-coated microchannels was assessed for CD8 T cells from 3 matched individuals (FAP01, FAP02, FAP07). **(E-F)** Migration through fibronectin-coated microchannels was assessed for CD8 T cells from 5 individual pairs (FAP01, FAP02, FAP03, FAP05, FAP07). (F) Left panel show sequential images of Hoechst-labelled CD8 T cells from a representative HD and matched FAP patient. 20x objective. Right panels show video microscopy frames (Video 1). 63x objective. Insets zoom in the front edge’s areas of migrating cells displaying cell lamellipodium (bracket) or lobe-shaped protrusions or blebs (empty arrowheads). Scale bars 10 µm.

To investigate the potential consequences of APC defects in a more physiological setting, we analyzed T cell chemotaxis through human umbilical vein endothelial cell (HUVEC) monolayers. For these experiments, we made use of T cells from three FAP patients and their matched healthy volunteers that had been conserved frozen and set back in culture. Cells from FAP patients displayed a diminished capacity to migrate through HUVEC monolayers (Figure 1B). Altogether, these data show that FAP T cells have a decreased capacity to perform chemotaxis towards CXCL12 through micropores or endothelial barriers.

We further studied FAP T cell migration at the single-cell level in the absence of chemokines in order to address whether this motility impairment is due to an intrinsic migration defect. We concentrated in the study of the CD8 T cell population since it was the most enriched upon activation. First, we measured migration of T cells in 3D collagen matrices, in which leukocytes rely mostly on cell contractility to move (17, 18). In this system, motility of CD8 T cells from FAP patients was not impaired with respect to matched healthy subjects (Figure 1C and Supplemental Figure 1B-C).

We next used microchannels in which cells migrate in a straight line, providing a simplified assay to study T cell locomotion (19, 20). Cells from FAP patients showed a significant increase in their migration when moving in collagen-coated microchannels (Figure 1D). Altogether, these data indicate that FAP CD8 T cells do not have intrinsic defects of their migration capacity in collagen-coated substrates.

Since T cell migration can be influenced by their adhesion to the extracellular matrix, we performed migration experiments in microchannels coated with fibronectin, which binds to VLA-4 at the T cell surface. Notably, migration speed of FAP CD8 T cells was significantly diminished under this condition (Figure 1E-F). In addition, these cells appeared more elongated and progressed emitting multiple bleb-like protrusions at the leading edge, while control cells displayed a larger front lamellipodium, indicating different modes of migration on fibronectin-coated surfaces (Figure 1F, Video 1).

Altogether, these data provide the first evidence that APC contributes to regulate T cell migration in confined microenvironments, as often found in tissues. The impact of APC depends on the composition of the extracellular matrix, indicating an adhesion-dependent phenomenon.

### VLA-4-mediated adhesion is impaired in T cells from FAP patients

The importance of adhesion to fibronectin to distinguish control from APC mutant cells, prompted us to analyze T cell adhesion forces to the fibronectin-interacting integrin VLA-4, using shear flow microchambers. To prevent heterogeneity of fibronectin coating, we used the cellular ligand for VLA-4, VCAM-1, in the presence of CXCL12 chemokine that enhances integrin-mediated adhesion (21). When increasing shear force was applied to bound CD8 T cells, FAP patient’s T cells detached at lower force than those from healthy subjects (Figure 2A-C; Video 2), indicating that APC mutations result in decreased T cell adhesion to VCAM-1.

**Figure 2:**
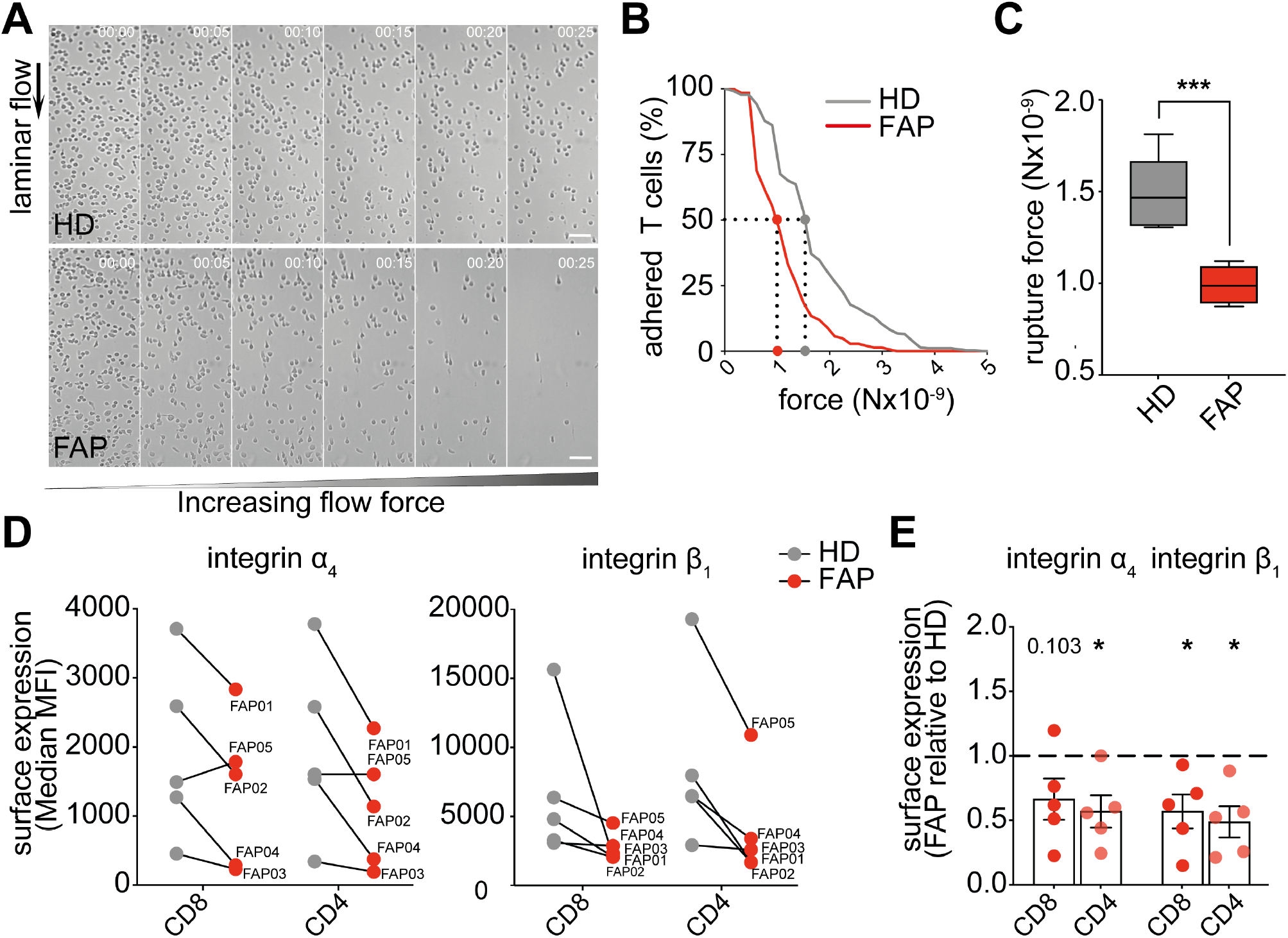
T lymphocytes from FAP patients display altered integrin-mediated adhesion. Adhesion strength of T cells from FAP patients (FAP) and their matched healthy donors (HD) to VCAM-1+CXCL 12-coated surface was assessed by shearing flow **(A-C)**, and surface expression of the α_4_ and β_1_ integrin VLA-4 (ligand for VCAM-1) in these cells was analyzed by flow cytometry **(D-E). (A-C)** CDS T cells were seeded in a laminar fiow chamber and submitted to increasing flow rates. The number of adherent cells per frame was counted and the flow rate at rupture was analyzed. Sequential images of detaching CDS T cells from a representative pair of HD and FAP (FAP05) are shown in **(A)** (Video 2). Flow’s direction is indicated. 10x objective. Scale bars 50 µm. **(B)** The number of adherent cells per frame was plotted for this representative individual pair as a function of the force. Dotted lines mark the rupture force necessary for detaching 50% of the cells from the substrate. **(C)** Boxes represent the rupture force of adhesion to VCAM-1+CXCL12-coated surfaces of T cells from 7 FAP patients and matched healthy individuals (FAP01 to FAP07). Statistical differences were calculated by Mann-Whitney unpaired test. *** p< 0.001. **(D-E)** Surface expression of α_4_ and β_1_ integrin subunits was measured by flow cytometry in 5 subjects’ pairs (FAP01 to FAP05). **(D)** Linked dots show the median fluorescence intensity of VLA-4 integrin α_4_ and β_1_ subunits for each FAP patient and its matched HD. **(E)** The VLA-4 median fluorescence intensity of each FAP patient was normalized to the one of his matched HD. Each dot represents 1 FAP patient and the dashed line shows the reference value of HD (normalized to 1). Statistical differences were calculated by one-sample two-tailed t-test. P-values are indicated or replaced by * when p< 0.05.

Consistent with their reduced adhesion to VCAM-1, the α_4_ and β_1_ subunits of VLA-4 were reduced in both CD8 and CD4 T lymphocytes from 4 out of 5 FAP patients tested (Figure 2D-E). Indeed, the relative expression of both proteins, calculated as the ratio of their median fluorescence intensity in FAP vs matched control cells, was significantly lower in the former (Figure 2E). One patient (FAP05) showed no reduction in α_4_ compared to its matched HD, yet it had a lower expression of the β_1_ subunit, which would reduce the number of integrin complexes and may affect the adhesion in flow chambers (Figure 2A-B).

Therefore, T cells from FAP patients present impaired VLA-4 integrin-mediated adhesion to VCAM-1, also displaying reduced expression of the VLA-4, potentially accounting for adhesion defects.

### Specific inhibition of APC expression in CEM T cells induces adhesion and migration defects similar to those observed in FAP patients’ T cells

FAP patients are heterozygous and carry a variety of APC mutations that may result in null or truncated APC protein expression of the mutated allele and normal expression of the wild-type allele. Moreover, the number of cells obtained from patients is limited, precluding certain types of experiments. In order to generate an experimental system with a more homogeneous and controlled APC defect, providing enough cells to analyze the underlying molecular and cellular mechanisms, we investigated the effects of silencing APC expression by small interfering RNA (siRNA) in CEM tumor T cells (22) (Figure 3A).

**Figure 3:**
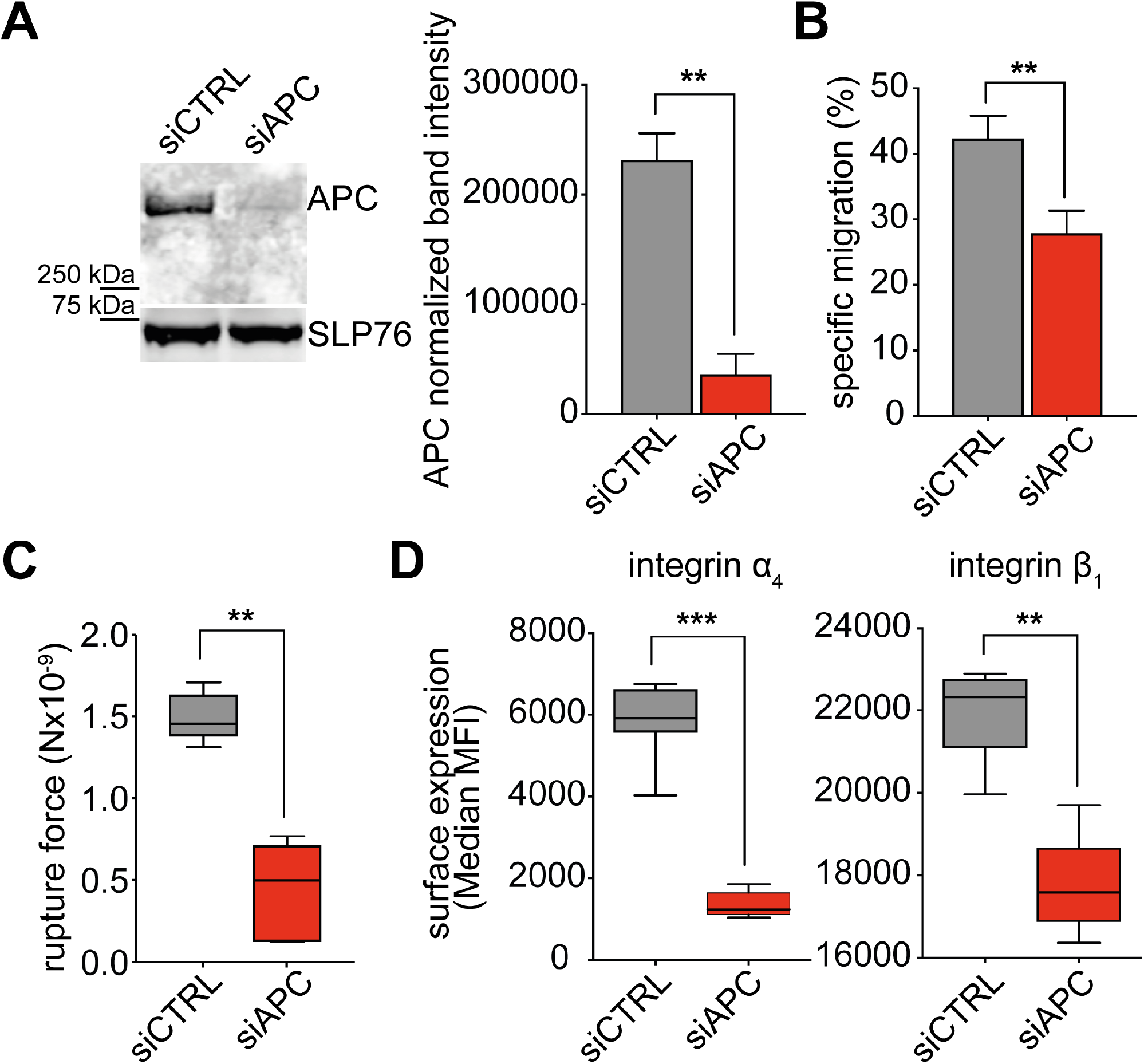
APC silencing impairs migration and adhesion of CEM T cells. CEM T cells were transfected with control (siCTRL) or APC (siAPC) siRNA oligonucleotides and used 72h after transfection. **(A)** Western blot showing the expression level of APC protein in siCTRL- and siAPC-transfected CEM T cells. A representative result is shown in left panel. Quantification of APC band intensity from 4 independent immunoblots, normalized by the intensity of SLP76 in the same sample, is shown in right panel. **(B)** Transmigration through transwell filters was analyzed for control and APC-silenced CEM T cells. Bar plots represent the mean + SD of T cell specific migration. Statistical differences (4 independent experiments) were calculated by Mann-Whitney unpaired test. **p< 0.01. **(C)** Control (siCTRL) and APC-silenced (siAPC) CEM T cells were seeded in VCAM-1+ CXCL12-coated chambers and a laminar shear flow of PBS was applied through the chamber (Video 3). Scale bars 50µm. Boxes display the rupture force from 4 independent experiments. Statistical differences were calculated by Mann-Whitney unpaired test. ** p< 0.01. **(D)** Cell surface expression of α_4_ and β_1_ integrin subunits was analyzed by flow cytometry. Boxes display the median fluorescence intensity of 4 independent experiments. Statistical differences were calculated by Mann-Whitney unpaired test. **p<0.01, ***p< 0.001.

In line with the results from FAP patients (Figure 1A and 2A-B), we found that CEM T cell ability to migrate through transwell filters in response to CXCL12 was impaired by APC-silencing (Figure 3B). Likewise, their adhesion to VCAM-1 + CXCL12-coated microchambers under shear flow was reduced (Figure 3C; Video 3). Finally, VLA-4 expression was reduced (Figure 3D), with α_4_ subunit being more affected than β_1_ (75% and 20% reduction, respectively).

### APC-silenced CEM T cells display altered substrate contact patterns

To further characterize the impaired adhesion and migration of APC-silenced cells, we analyzed the interaction of migrating cells with VCAM-1 + CXCL12-coated surfaces, imaging the area of cell-substrate contact by Interference Reflection Microscopy (IRM) (14, 23). APC silencing changed cell shape, resulting in increased contact area, but weaker engagement with the surface, as shown by lower IRM intensity per cell (Figure 4A-C), in line with the reduced adhesion to VCAM-1 and the lower VLA-4 integrin expression (Figure 3C-D). Furthermore, APC-silenced T cells more often displayed rounder contact shapes (Figure 4A,C) with lobe-shaped protrusions (Figure 4A, empty arrowheads), while control cells were more elongated with long, thin, IRM/F-actin/VLA-4-positive adhesive filopodia (Figure 4A, arrowheads, C-D).

**Figure 4:**
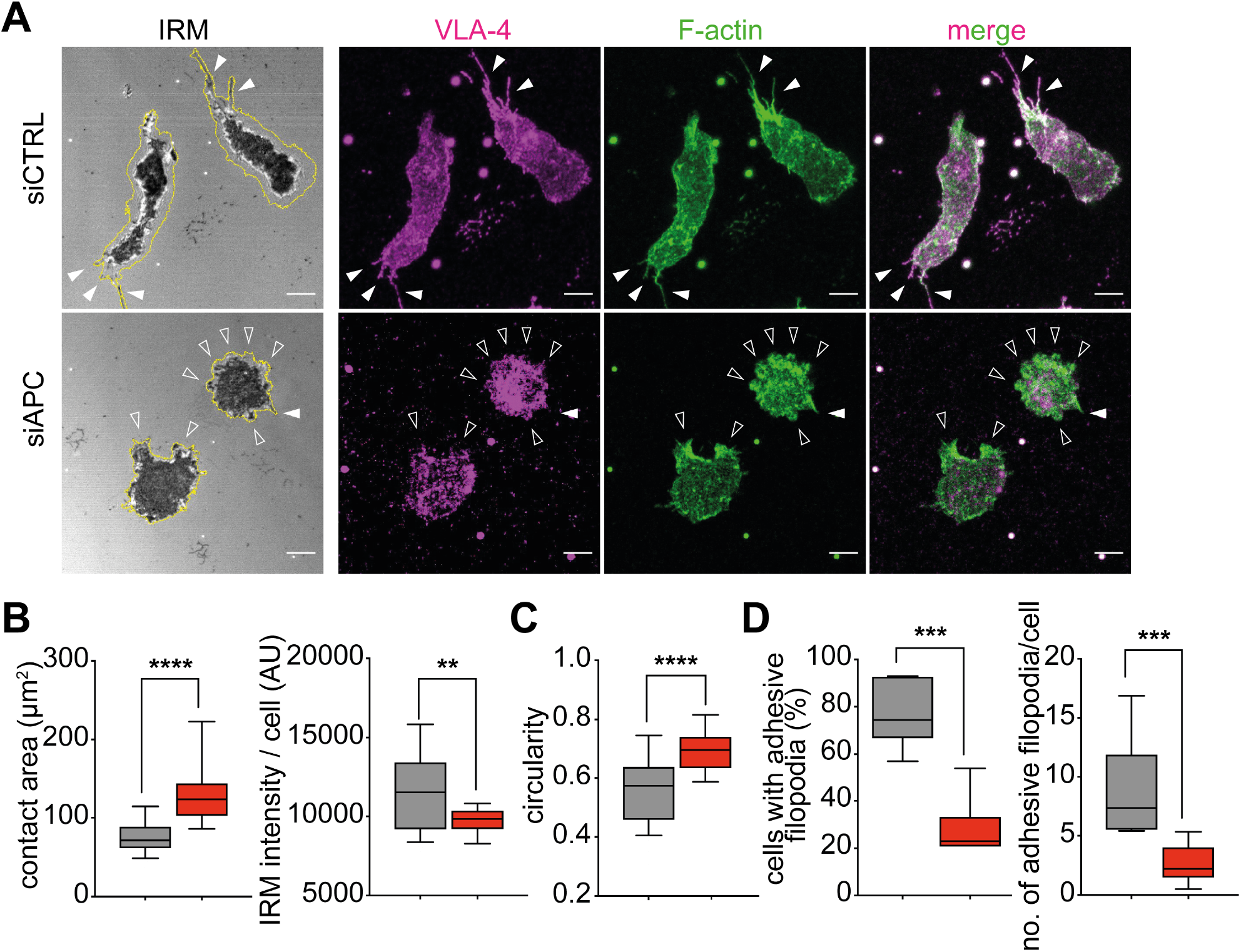
APC silencing alters VLA-4-dependent cell adhesion patterns. **(A-D)** CEM T cells were transfected with control (siCTRL) or APC (siAPC) siRNA oligonucleotides. Three days later, cells were seeded on VCAM-1+CX-CL12-coated glass dishes and allowed to adhere and migrate during 10 minutes. Thereafter, non-adherent cells were washed out and dishes were fixed and stained for VLA-4 and F-actin. T cell adhesive contacts of representative siCTRL- and siAPC-transfected cells analyzed by IRM and fluorescence confocal microscopy are shown. **(A)** Cell were segmented by thresholding on the F-actin channel to define their outline (yellow line). Darkest areas in the IRM image correspond to areas where the cell attaches the substratum (left panel) and reveals a stronger adhesion. siCTRL T cells adhere tightly to the surface and display lateral and back thin protrusions (filled arrowheads) contacting the substratum. These protrusions are enriched in VLA-4 and F-actin, as displayed in the immunofluorescence images. Empty arrowheads indicate lobe-shaped protrusions typical of siAPC T cells. 63x objective. Scale bars 5µm. **(B-D)** Boxes display measurements from 4 independent experiments (n>100). Statistical differences were calculated by Mann-Whitney unpaired test. **** p< 0.0001, *** p< 0.001, ** p< 0.01. **(B)** T cell contact area with the substrate (in µm^2^; left box blot) and total pixel intensity of the contact area (in arbitrary units [AU]; right box plot) were calculated on IRM images. **(C)** Cell circularity was measured on IRM images with the following formula: circularity= 4π*area/perimeter^2^. A circularity value of 1.0 indicates a perfect circle. As the value approaches 0.0, it indicates an increasingly elongated shape. **(D)** Percentage of cells displaying VLA-4-enriched filopodia (left box plot) and number of filopodia per cell (right box plot) were measured on fluorescence images.

These data add evidence that APC plays a crucial role in regulating adhesion and cytoskeleton coordination, necessary for lymphocytes to properly migrate on adhesive substrates.

### APC-silenced CEM T cells migrate processively extending and retracting unstructured multiple lobes

In agreement with the phenotype highlighted by IRM analyses, imaging live cell dynamics on the same surface showed APC-silenced CEM T cells often displaying several pseudopodia and round protrusions (Figure 5A, empty arrowheads), whereas control cells rather displayed elongated shapes, with an evident front edge led by a dominant lamellipodium (Figure 5A, brackets; see also Video 4).

**Figure 5:**
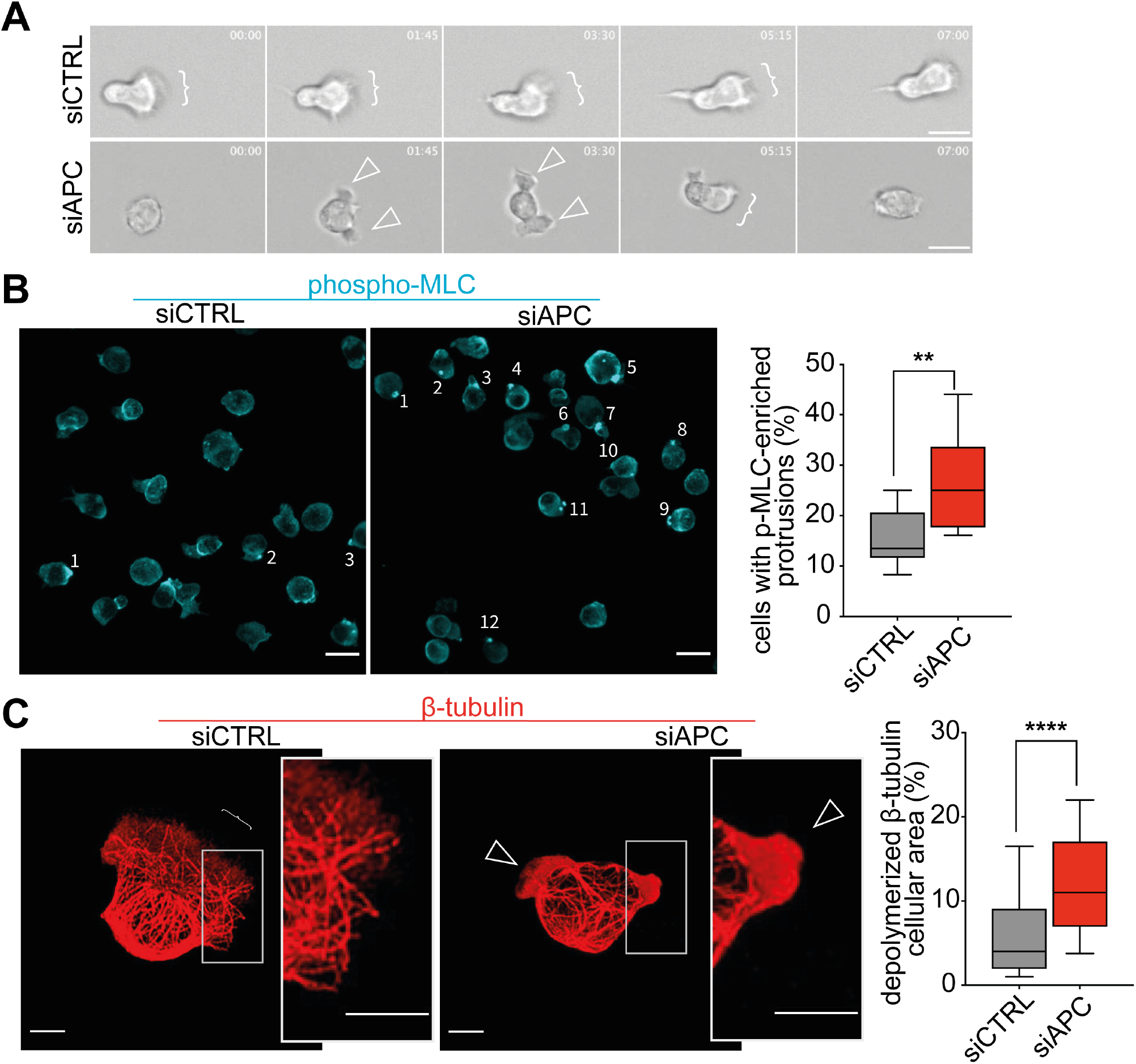
APC silencing modifies phospho-myosin and microtubule pattern in T cell protrusions. CEM T cells were transfected with control (siCTRL) or APC (siAPC) siRNA oligonucleotides and used after 72h. Cells were seeded on VCAM-1+CXCL 12-coated glass dishes **(A)** or coverslips **(B, C)** and allowed to adhere and migrate during 10 minutes. Thereafter, non-adherent cells were washed out. **(A)** The dynamics of migrating cells was analyzed by live cell microcopy. Sequential frames of representative control and APC-silenced migrating T cell are shown (Video 4). Brackets indicate cell lamellipodia, empty arrowheads point at pseudopodia. 20x objective. Scale bars 10 µm.**(B, C)** Migrating cells were fixed and stained using anti-pMLC or anti-β-tubulin antibodies, respectively. **(B)** Numbered cells in immunofluorescence images show accumulation of pMLC in small membrane protrusions. 63x objective. Scale bars 5µm. The percentage of cells with this particular pMLC pattern was measured in 3 independent experiments (n>100) and represented as box plot on the right. Statistical differences were calculated by Mann-Whitney unpaired test. **p< 0.01. **(C)** lmmunofluorescence images show microtubule network in siCTRL and siAPC CEM T cells. Insets show enlarged images of cell lamellipodium (bracket) or membrane extensions containing depolymerized β-tubulin (empty arrowheads). 63x objective. Scale bars 5µm. The percentage of cell area displaying depolymerized β-tubulin was calculated in 3 independent experiments (>100) and represented as box plot. Statistical differences were calculated by Mann-Whitney unpaired test. ****p< 0.0001.

The dynamic behavior of APC-silenced T cells characterized by consecutive extension and retraction of pseudopodia prompted us to test whether these events may be acto-myosin-driven (24). Thus, we looked for the presence of phosphorylated myosin light chain (p-MLC) and observed phospho-MLC-enriched small protrusions more often present in APC-silenced T cells (Figure 5B).

Finally, more accurate analysis of membrane extensions showed that APC-silenced T cells more often had protrusions containing depolymerized tubulin (Figure 5C, empty arrowheads and zoomed area), instead of radial microtubules pointing to the front edge as in control cells (Figure 5C, brackets and zoomed area).

Therefore, APC silencing affects microtubule cytoskeleton in lamellipodia, destabilizing lamellipodium areas and favoring lobe-type membrane extensions.

### APC silencing correlates with reduced ERM phosphorylation and T cell cortical rigidity

To explore the molecular bases of increased bleb-type protrusions in APC-silenced cells, we investigated whether ERM phosphorylation and cortical rigidity were affected by APC silencing. Indeed, APC, through its interaction with Dlg1, may be part of a complex with ERMs that controls microtubule network organization at the T cell cortex (6, 25). Moreover, ERMs, through their Thr phosphorylation control the interactions between plasma membrane and the cortical actin cytoskeleton thus regulating cortical rigidity and bleb generation (26). Additionally, ERMs control Rho signaling and may therefore regulate local myosin-mediated contractility and keep the balance between lamellipodial *versus* bleb-type protrusions (27, 28). Hence, we evaluated cell deformability under centrifugal force and ERM phosphorylation (29, 30). APC silenced cells showed a significantly higher tendency to flatten than control cells, as assessed by their increased aspect ratio and deformability index (Figure 6A-B). Moreover, APC silenced cells displayed lower levels of pERMs (Figure 6C), which may result in impaired cell cortex organization and stability (26, 30).

**Figure 6.**
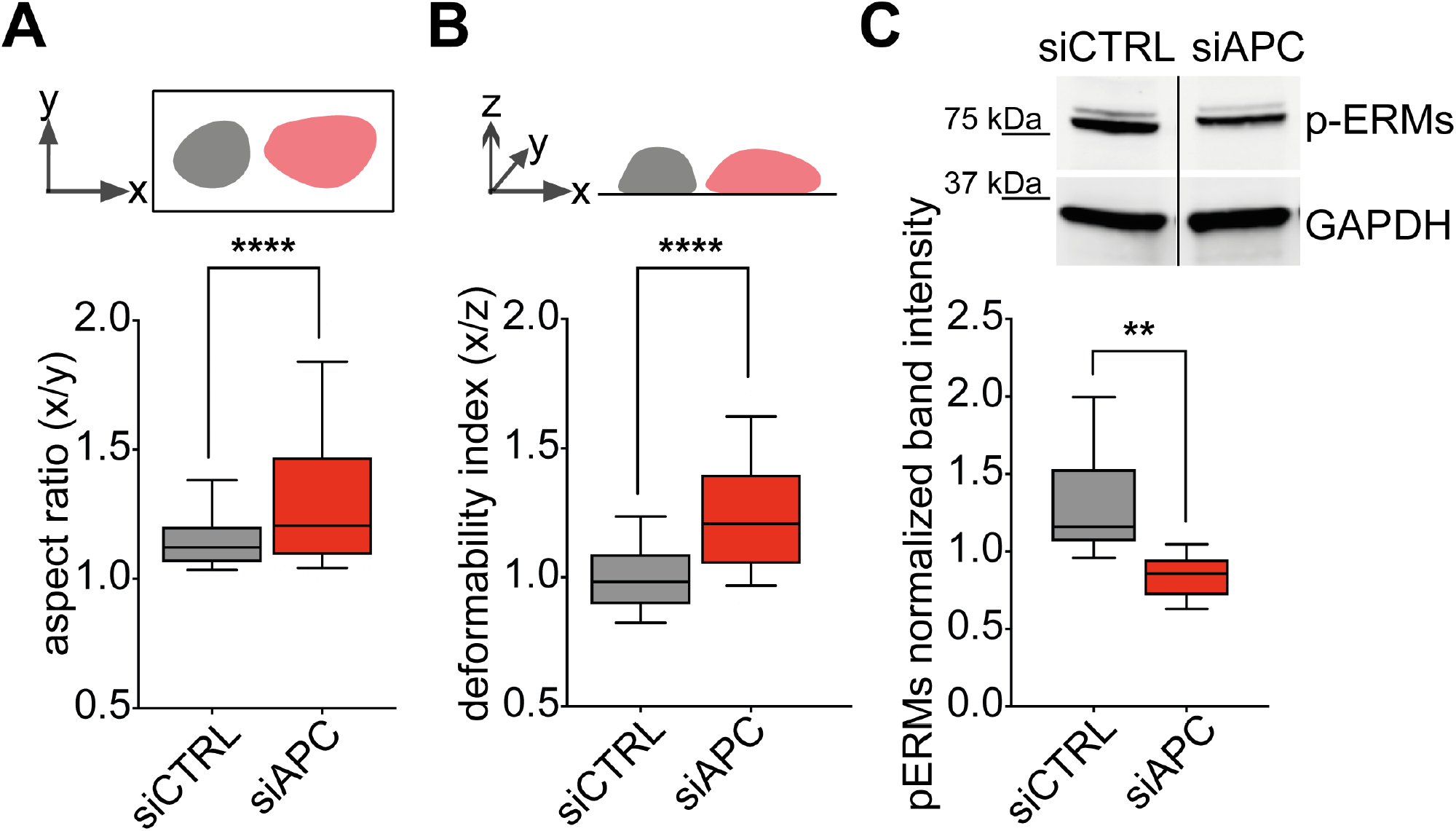
APC silencing alters cortical rigidity and ERM phosphorylation. CEM T cells were transfected with control (siCTRL) or APC (siAPC) siRNA oligonucleotides and used 72h after for the following analyses. **(A**,**B)** Cell-Trace Far Red dye-labeled cells were settled on poly-L-lysine-coated coverslips. After adding a paraformaldehyde solution, cells were centrifuged at 3,724×g for 10 minutes. Cell deformation was quantified for each cell by measuring the aspect ratio **(A)** and the deformability index **(B)**, and represented as box plots. Statistical differences were calculated by Mann-Whitney unpaired test. ****p< 0.0001. Aspect ratio and deformability indexes were calculated as follows: major axis (x)/minor axis (y) and major axis (x)/thickness (z), respectively. The more values deviate from 1, the more elongated or deformed cell shapes are. **(C)** Phospho-Ezrin, Thr567, higher band and phospho-Moesin, Thr558, lower band and GAPDH levels were assessed by western blotting in siCTRL- and siAPC-transfected CEM T cells. ERM phosphorylation was quantified by near-fluorescence intensity and normalized to GAPDH intensity in the same sample. A representative immunoblot is shown in left panel. Lanes were run on the same gel but were noncontiguous. Data are mean + SEM of 4 independent experiments. Statistical differences were calculated by Mann-Whitney test. **p<0.01.

Therefore, APC silencing alters the organization of actin and microtubule cytoskeleton leading to reduced cortical rigidity. Consequently, APC-silenced T cells appear more prone to form several membrane extensions instead of a dominant lamellipodium.

## Discussion

Migration is a crucial step for immune functions, including T cell anti-tumor responses. We show here that the tumor suppressor and cell polarity regulator APC is key for T cell adhesion and migration. Indeed, our data reveal that T cells from FAP patients carrying APC mutations and APC-silenced CEM T cells display impaired migration. We identified integrin-mediated cell adhesion and cytoskeleton organization and stability as features targeted by APC defects. These are key requirements for T cells to migrate through physiological environments, as endothelium and lymphoid organs, or tumor tissues (31, 32).

Cells migrate as the result of the interplay between physical constraints (e.g. stiffness, porosity, order), substrate interactions (e.g. integrin-mediated adhesion) and chemical signals (e.g. chemokines) from the local microenvironment. These are mechanically integrated into membrane and cytoskeleton-mediated cellular processes, such as adhesion, traction, protrusion, deformation, and polarization (11, 33).

Using calibrated tools for *in vitro* cell migration, we identified APC as key for chemokine-induced T cell migration through micropores and HUVEC monolayers, and for spontaneous migration through narrow adhesive microchannels. Patients’ T cells showed impaired migration when migrating within microchannels when coated with fibronectin, an extracellular matrix VLA-4 ligand. In contrast, on collagen, whose binding is VLA4-independent, FAP patients’ T cells migrated faster than those from healthy subjects. Similarly, migration in 3D collagen matrices was not impaired in T cells from FAP patients. Therefore, APC mutations result in impaired VLA-4-dependent migration in confined environments.

In line with these findings, APC defects impaired adhesion forces on surfaces coated with the VLA-4 integrin ligand VCAM-1 and the chemokine CXCL12, whose signaling reinforces integrin-mediated adhesion. Interestingly, most of the FAP patients displayed a reduced VLA-4 cell surface expression in *ex vivo* activated T cells. Although the molecular mechanism linking APC defects to integrin expression impairment is unknown, this could be, at least in part, a cause of defective adhesion in patients’ T cells.

APC mutations in FAP patients may result in differential functional phenotypes. Moreover, being a rare disease, the number and volume of samples was limited and precluded some mechanistic analyses. Therefore, to specifically target APC, enhance the phenotype and gain access to larger number of cells, we investigated the effect of APC silencing in CEM leukemia T cells, a suitable model for T cell migration analyses (22). APC-silenced CEM T cells phenocopied migration, adhesion and VLA-4 expression defects observed in FAP patient’s T cells, supporting the APC involvement in these functions.

To gain insight into the molecular and cellular processes involved, we used a 2D migration set up, where cells were allowed to migrate on VCAM-1 and CXCL12-coated surfaces then fixed and analyzed by microscopy. While control cells adopted elongated shapes with distinguishable uropod and front lamellipodium, and frequent F-actin/VLA-4-enriched filopodia, APC-silenced cells were rounder and with less filopodia. Additionally, they rarely polarized, rather they extended and retracted multiple pseudopodia on different directions, possibly affecting their migration persistence (34). Generation of multiple pseudopodia was reminiscent of migration defects in T cells overexpressing a PKCζ kinase-dead mutant (35). Interestingly, Cdc42 and Par6-PKCζ regulate the association of Dlg1 and APC to control cell polarization in migrating astrocytes (36).

Consistent with adhesion force differences, IRM revealed that control cell contacts with the substrate were tighter, although covering a smaller surface. APC-silenced T cells displayed also multiple small protrusions enriched in phosphorylated-myosin and pseudopodia lacking structured microtubules. Since microtubule disorganization may trigger acto-myosin-driven retraction, both observations could be related (37, 38). In contrast, in control cells, microtubules appeared structured, reaching lamellipodial edges and pMLC-enriched protrusions were less frequent. This may facilitate sustained front edge advancing (39).

APC is part of the same polarity complex as Dlg1, that interacts with ezrin end moesin, which in turn control cell cortex rigidity (25, 30, 40, 41). We found that APC silencing reduced T cell cortical rigidity and steady state phosphorylation of ezrin and moesin, suggesting cytoskeleton relaxation in APC-silenced T cells (30). This could also influence adhesion by altering integrin-cytoskeleton anchoring properties that control integrin avidity (42, 43). Finally, because of looser membrane interaction with the cortical cytoskeleton in APC-deficient cells, contraction forces and the consequent increase in hydrostatic pressure may more easily favor membrane extensions and an ameboid-like migration on adhesive substrates (26, 44). Interestingly, MLC phosphorylation and ERM proteins have been proposed to drive bleb-based motility of sphingosine-1-phosphate stimulated T cells (44). Our findings suggest that similar mechanisms and molecular actors may also regulate bleb-type extensions of FAP patients’ T cells in fibronectin-coated microchannels. Indeed, bleb-mediated migration has been reported to preferentially occur under certain physically confined conditions (26, 45).

In conclusion, our data provide the first evidence that APC orchestrates the coupling of cytoskeletal-mediated forces and VLA-4-dependent adhesion necessary for T cell migration as physiologically occurs across endothelial barriers, lymphoid organs, inflamed tissue and tumors. Consequently, APC defects may impair immune surveillance and effector functions of T cells, thus favoring tumorigenesis initiated by defects in epithelial cell differentiation in FAP patients.

## Methods

### Patients

Seven FAP patients and their respective sex-, age- and ethnicity-matched healthy donors were recruited through the *Association Polyposes Familiales France* and the Institut Pasteur ICAReB core facility (NSF 96-900 certified, from sampling to distribution, reference BB-0033-00062/ICAReB platform/Institut Pasteur, Paris, France/BBMRI AO203/1 distribution/access: 2016, May 19th, [BIORESOURCE]), under CoSImmGEn protocols approved by the Committee of Protection of Persons, Ile de France-1 (No 2010-dec-12483 for healthy donors and No 2018-mai-14852 for FAP patients). Informed consent was obtained from all donors. Subjects were 4 women and 3 men. Average age was 48.8 years for healthy donors (range 29-63) and 47.7 years for patients (29-60). Age matching was within a 0-5 years difference (average 2.6). APC mutations included frameshift mutations potentially leading to lack of protein expression of the mutated allele or expression of truncated forms.

### Cells and transfections

Peripheral blood mononuclear cells were purified from FAP patients or healthy donors by Ficoll-Hypaque centrifugation and activated with TransAct (Miltenyi-Biotec 130-111-160) (1:100) and recombinant human IL-2 (100 U/mL; PeproTech) in RPMI-1640 medium supplemented with GlutaMAX-I (Gibco), 5% human serum, 1 mM sodium pyruvate, nonessential amino acids, 10 mM HEPES, 1% penicillin-streptomycin (v/v). After 5 days, CD4 T cells were purified using positive magnetic sorting (Miltenyi-Biotec 130-045-101) and CD8 T cells recovered from the non-retained population. Aliquots of activated T cells were frozen and settled back in culture later on for further experiments.

CEM T cells (acute lymphoblastoid leukemia cell line; ref.46) were cultured in RPMI-1640 medium supplemented with 10% fetal bovine serum (FBS) and 1% penicillin-streptomycin (v/v).

For siRNA transfections, 1nmol of control or APC siRNA was used per 10^7^ CEM T cells. Sequences used were previously described (15, 16). Two transfections (1400 V, 10 ms, 3 pulses) were performed at 24h interval with a Neon Transfection System (Invitrogen). Cells were analyzed 72h after.

### Chemotaxis

Primary T cells or CEM T cells (1×10^6^ cells/mL in RPMI-1640, 0.5% BSA) were deposited in the upper inserts of transwell wells (Nunc). For trans-endothelial chemotaxis, a HUVEC monolayer was previously grown on these inserts as follows. Confluent HUVEC cells were gently trypsinized and seeded (0.2×10^6^ in 300μL of complete EBM2 medium) in the upper compartments of the transwell inserts, previously coated with serum at 37°C for 2h. The lower compartments contained the same medium, were refreshed the following day. After 3 days of culture, the integrity of confluent HUVEC monolayer was assessed by microscopy observation (Supplemental Figure 1A) and by measuring the permeability of the monolayer using FITC-dextran (Sigma #FD20S) diffusion. For T cell trans-endothelial chemotaxis, the upper compartments were gently rinsed in warm 0.5% BSA RPMI medium before seeding T cell.

The lower chambers contained the same medium supplemented or not with different concentrations of recombinant CXCL12 (R&D 350-NS) (40 ng/mL for CEM T cells if not otherwise indicated). After, 90 min and 60 min of incubation, respectively for primary and CEM T cells, at 37°C in 5% CO_2_, inserts were removed and the same volume of migrated and input populations was analyzed by flow cytometry. The percentage of CD8/CD4 T cells was assessed for primary T cells.

### Migration in microchannels

Micro-channel experiments were performed as described(47). Briefly, customized polydimethylsiloxane (PDMS) micro-chips were coated with fibronectin (10 µg/mL) (Sigma-F1141) or collagen type I (SureCoat™-5057) for 1h at room temperature (RT), then washed with PBS. T cells were labeled with Hoechst (Thermo-Fisher-Scientific-33342) (200 ng/mL) 30 min at 37°C, washed twice and loaded in the micro-chips. Cell migration at 37°C in 5% CO_2_ was recorded overnight at 1 image/min with a Leica DMi8 video-microscope (10x/0.40 NA phase objective) and analyzed using a custom software.

### Cell migration in 3D collagen matrices

T cell migration in 3D was evaluated in customized PDMS chambers filled with a collagen I (Corning 354249) mix (4mg/mL) containing 0.72 ×10^6^ cells/mL. Gels were allowed to polymerize for 20 min at 37 °C, 5% CO_2_ and equilibrated in RPMI-1640 5% human serum. Cell migration at 37°C in 5% CO_2_ was recorded at 2 images/min (10x/0.40NA phase objective). and analyzed using a custom software.

### Rupture force in laminar flow chambers

Flow chambers (μ-slide, Ibidi) were coated with rhVCAM-1/Fc Chimera (R&D 862-VC) + recombinant CXCL12 (R&D 350-NS) (1μg/mL and 100 ng/mL in PBS, respectively) for 3 hrs at RT, rinsed and equilibrated with RPMI-1640 for 10 min at 37°C, then loaded with T cells for 10 min at 37°C. A flow rate of PBS (0 to 50 mL/min at 37°C) was applied through the temperature-controlled chamber for 92 sec using a computer-driven syringe pump (SP210iW, World Precision Instruments) synchronized with image acquisition (3 images/sec) using an inverted transmission microscope (Axio Observer D1; Zeiss, 10x/ NA 0.3 objective) and Micro-Manager software (48). Images were analyzed using Fiji software (49) and the Cell Counter plugin. Forces necessary for cells to detach were calculated by determining the flow rate at rupture as described (16).

### Flow cytometry

T cell surface protein staining was carried out during 40 minutes at 4°C with appropriate conjugated fluorophore antibodies (Table 2) prepared in PBS 2% serum 0.05% NaN. 100.000 events in the viability gate (Fixable Viability Stain 450, BD 562247) were acquired on a MACSQuant Analyzer (Miltenyi-Biotec) and processed with Kaluza software.

### Fluorescence and Interference Reflection Microscopy (IRM)

Coverslips or glass-bottom microwell dishes (MatTek) were coated with rhVCAM-1/Fc Chimera (R&D 862-VC) + recombinant CXCL12 (R&D 350-NS) (respectively 1μg/mL and 100 ng/mL in PBS) 2 hours at 37°C. They were further washed and blocked 30 minutes with PBS 1% BSA, CaCl_2_ and MgCl_2_. CEM T cells (10^5^), resuspended in adhesion medium (PBS 1% FBS, 0.9 mM CaCl_2_, 0.5 mM MgCl_2_ supplemented with HEPES), were allowed to migrate on coverslips or dishes for 10 min at 37°C, 5% CO_2_. Non-adhering cells were washed out and the remaining ones were fixed 12 min in 4% PFA, blocked in PBS 1% BSA over night at 4°C. For microtubule detection, coverslips were additionally incubated 5 min at −20°C in methanol before blocking. Fixed samples were incubated 1 hour at RT with PBS 1% BSA, 0.1% Triton X100 containing primary Abs (Table 1), gently washed in PBS 1% BSA and incubated 45 min at RT with the corresponding fluorochrome-coupled secondary Abs and phalloidin (Table I). Coverslips were mounted on microscope slides using ProLong Gold Antifade mounting medium with DAPI (Life-Technologies).

**Table 1.** Primary and secondary antibodies / reagents used for immunofluorescence.

| Antibodies/reagents –Clone | Host-Isotype | Reference | Dilution |
| --- | --- | --- | --- |
| Acti-stain 488 Fluorescent Phalloidin | Phallotoxin | Cytoskeleton PDHG1 | 1:50 |
| Anti-human CD49D purified clone 9F10 | Mouse IgG1 | eBioscience 14-0499 | 1:75 |
| Anti-human CD29 purified clone TS2/16 | Mouse IgG1 | eBioscience 14-0299 | 1:75 |
| Anti-Phospho-Myosin Light Chain 2 (Ser19) | Mouse IgG1 | Cell Signaling Technology 3675 | 1:100 |
| Anti-tubulin, beta, clone KMX1 | Mouse IgG2b | Millipore MAB3408 | 1:500 |
| Anti-Mouse IgG2b, Human ads-FITC | Goat | SouthernBiotech 1090-02 | 1:100 |
| Cy3-conjugated AffiniPure Anti-Mouse IgG1 | Goat | Jackson ImmunoResearch 115-165-205 | 1:100 |
| Cy5-conjugated AffiniPure Anti-Mouse IgG1 | Goat | Jackson ImmunoResearch 115-175-205 | 1:100 |
| Anti-Fluorescein/Oregon Green, Alexa Fluor 488 conjugate | Goat IgG1 | Invitrogen A11096 | 1:100 |

**Table 2.** Antibodies used for flow cytometry.

| Antibodies –Clone | Host-Isotype | Reference | Dilution |
| --- | --- | --- | --- |
| PE/Cy5 anti-human CD3 clone HIT3a | Mouse IgG2a | BD 555341 | 1:30 |
| FITC anti-human CD4 clone RPA-T4 | Mouse IgG1 | Biolegend 300506 | 1:50 |
| APC/Cy7 anti-human CD8a clone RPA-T8 | Mouse IgG1 | Biolegend 301016 | 1:30 |
| PE anti-human CD49D clone MZ18-24A9 | Mouse IgG2b | Miltenyi Biotec 130-093-282 | 1:50 |
| PE/Cy7 anti-human CD29 clone TS2/16 | Mouse IgG1 | Biolegend 303026 | 1:50 |

Images were acquired with an LSM700 confocal microscope. For IRM, we used an LSM780 confocal microscope equipped with an 80/20 beam splitter and a variable bandwidth chromatic filter. Confocal optical sections (63x/1.40 NA objective) were acquired using ZEN software (Zeiss). Images were analyzed with Fiji software (49).

### Random migration on adhesive surfaces

Glass-bottom microwell dishes (MatTek) were coated as described above. CEM T cells (10^5^) were resuspended in adhesion medium and seeded on dishes 10 minutes at 37°C. Non-adhering cells were washed out and the dish was placed in the thermostatic humid chamber of a Definite-Focus video-microscope. Cell migration at 37°C in 5% CO_2_ atmosphere was recorded at 5 images/min during 20 minutes (20x/0.40 NA objective) and analyzed using Fiji software (49).

### Cortical rigidity

The experimental procedure has been described (29, 30). siRNA-transfected CEM T cells were labeled with 0.5 mM CellTrace Far Red DDAO-SE (Invitrogen C34553) and resuspended in RPMI-1640 without serum. Cells were settled on Poly-L-lysine coated coverslips and placed in 24-well plates. Same volume of 4% PFA was added to the wells, and plates were centrifuged 10 min at 3,724×g. Coverslips were then mounted on slides using ProLong Gold Antifade mounting medium. Images were acquired with a LSM700 confocal microscope (Zeiss) (63x/1.40 NA objective) and analyzed using Fiji software (49).

### Western blot

For immunodetection of APC and phosphorylated proteins, respectively, 2×10^6^ and 1×10^6^ of CEM T cell lysates were processed as previously described^14,15^. Primary antibodies (Table 3) were incubated overnight at 4°C. Secondary antibodies conjugated with AlexaFluor 680 or DyLight 800 (Thermo-Fisher-Scientific) were applied for 35 min at RT in the dark. Detection was performed using Odyssey Classic Near-Infrared Imaging System (LI-COR-Biosciences).

**Table 3.**
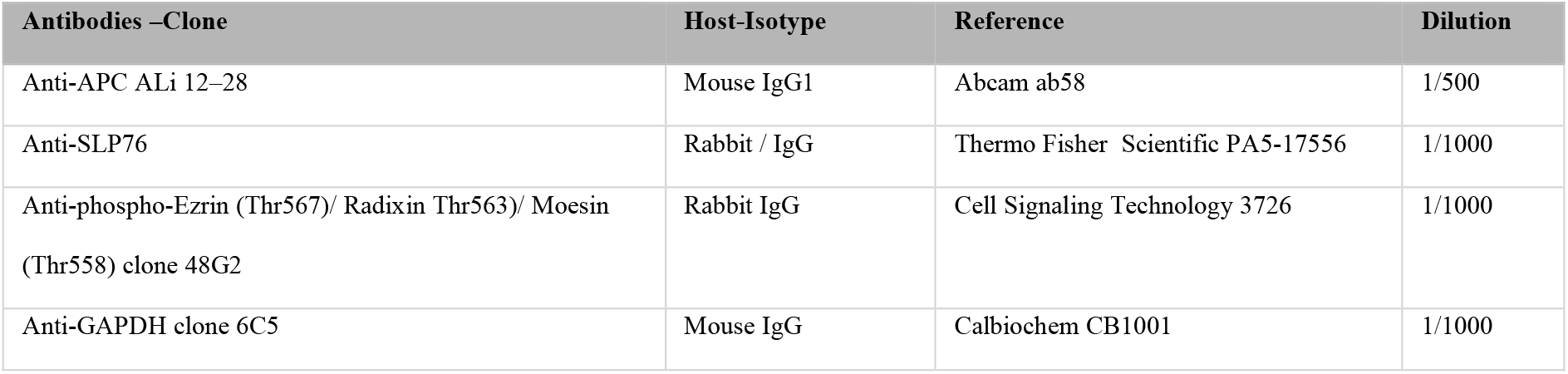
Antibodies used for western blot.

### Statistical analysis

Statistical analyses were carried out using GraphPad Prism V.9. Details are depicted in individual figure legends. The p values are represented as follows: ****p<0.0001, ***p<0.001, **p<0.01, *p<0.05, ns p≥0.05.

## Data Availability

All data will be made available upon request addressed to the corresponding authors.

## Author contributions

Contribution: M.M., A.A., V.D.B. conceived the project. M.M., P.V., T.R. designed and performed experiments, analyzed and interpreted data. M.M., C.C., M.J. prepared donors ‘samples and C.C. provided technical and laboratory management support. E.E. performed IRM imaging. H.L., C.R., M.N.U. provided access to patient and healthy donor samples. MM., A.A. and V.D.B. wrote the manuscript. J.D., P.V. provided expertise on T cell migration and critical manuscript reading.

All authors read and approved the final manuscript.

## Acknowledgements

We are in debt with the *Association Polyposes Familiales France* and the Institut Pasteur ICAReB core facility for their key contribution to the recruitment of FAP patients and healthy volunteers. We deeply thank all the volunteers for their participation in this project. We are grateful to Drs Sandrine Etienne-Manneville, Emmanuel Donnadieu and Fernando Sepulveda for scientific advice. We kindly thank Daria Bonazzi for expert assistance with HUVEC cells. We gratefully acknowledge the technical support provided by Julien Fernandes and Audrey Salles and the UtechS Photonic BioImaging at Institut Pasteur.

This work was supported by grants from *La Ligue Nationale contre le Cancer, Equipe Labellisée Ligue 2018*, and institutional grants from the Institut Pasteur and INSERM. UtechS Photonic BioImaging is supported by the National Research Agency (France BioImaging; ANR-10–INBS–04; Investments for the Future). M.M. is a scholar of the Pasteur Paris University International Doctoral Program, supported by the Institut Pasteur and the European Union Horizon 2020 Research and Innovation Programme under the Marie Sklodowska-Curie grant agreement 665807 (COFUND-PASTEURDOC). MM is currently funded by “Allocation doctorales 4ème année de thèse-La Ligue Contre Le Cancer”.

