## Supplemental Material for "The tumor suppressor Adenomatous polyposis coli regulates T lymphocyte migration. Insights from familial polyposis patients"

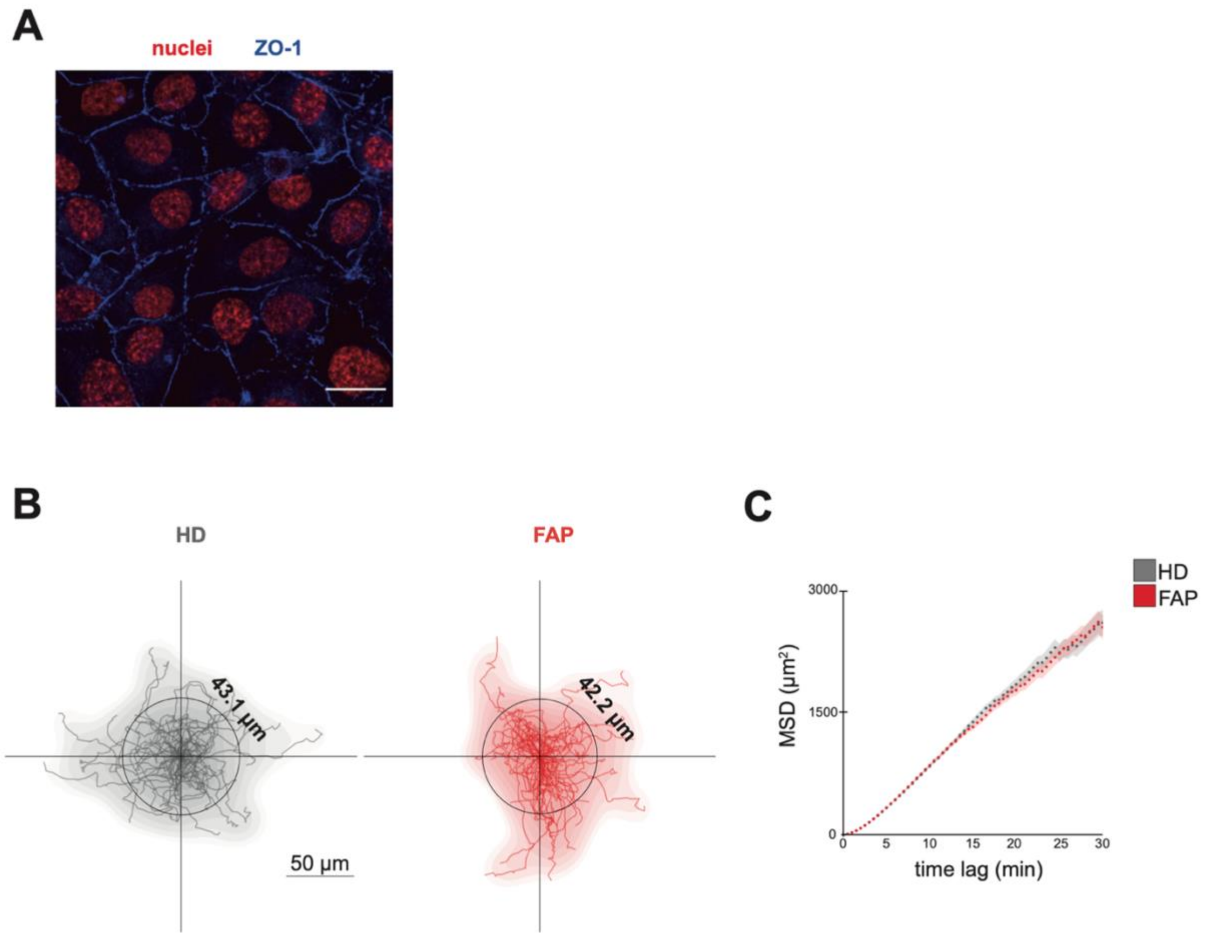


**Supplemental Figure 1: Assessment of HUVEC cell layers for T cell trans-endothelial chemotaxis and analysis of migration in collagen matrices (Related to Figure 1B-C)**

(A) To assess trans-endothelial chemotaxis, HUVEC cells were seeded on serum-coated 5 µm-por transwell filters. Three days later, the integrity of confluent HUVEC monolayer was analyzed by fluorescence microscopy. ZO-1 staining (blue) indicates mature tight junctions between HUVEC cells in the layer. Cell nuclei are stained in red. 63x objective. Scale bar 20 µm.

(B) CD8 T cells were assessed for 3D migration in collagen gels. Trajectory plots of CD8 T cells migrating in 3D collagen gels are shown for one representative individual pair (FAP03) out of four. Migration was imaged during 30 minutes. The starting point of each trajectory was translated to the origin of the plot.

(C) Mean squared displacement quantified from the data depicted in (B).

**Legends for videos**

**Video 1: FAP patients' CD8 T cells display impaired migration in fibronectin-coated microchannels (Related to Figure 1E-F)**. CD8 T cells from a HD or FAP were allowed to migrate in microchannels of 4µm width and 5µm height, previously coated with fibronectin. HD T cell migration (top panel) is faster and driven by a front dominant lamellipodium, whereas the FAP T cell one (bottom panel) is slower and characterized by the formation of blebs. This video is representative of 5 HD-FAP individual pairs. Time stamp is in hr:min:sec. 63x oil objective. Scale bar 10µm.

**Video 2: FAP patients' CD8 T cells detach at lower laminar flow forces in adhesion chambers (Related to Figure 2A-B).** CD8 T cells from a HD or FAP are allowed to seed in a temperature-controlled flow chamber, previously coated with VCAM-1 + CXCL12. A linear flow rate ramp of PBS (37°C), increasing from 0 to 50 mL/min, is applied to the chamber for 92s using a syringe pump. FAP patients' T cells (right panel) detach at lower flow intensity than HD cells (left panel), indicating defects in their adhesion strength. This video is representative of 7 HD-FAP individual pairs. Time stamp is in min:sec. 10x/0.3 objective. Scale bar 50µm.

**Video 3: APC-silenced T cells detach at lower laminar flow forces in adhesion chambers (Related to Figure 3C)** siRNA-transfected CEM T cells are allowed to seed in a temperature-controlled flow chamber, previously coated with VCAM-1 + CXCL12. Image acquisition starts when a linear flow rate ramp of PBS (37°C), increasing from 0 to 50 mL/min, is applied to the chamber for 92s using a syringe pump. siAPC T cells (right panel) detach at lower flow intensity than siCTR cells (left panel), indicating defects in their adhesion strength. This video is representative of 4 independent experiments. Time stamp is in min:sec. 10x/0.3 objective. Scale bar 50µm.

**Video 4: APC-silenced CEM T cells display a pseudopodia extension-retraction migration mode when migrating on adhesive substrates (Related to Figure 5A)**. siRNA-transfected CEM T cells are allowed to migrate on VCAM-1 + CXLC12-coated dishes. siCTR T cell (left panel) migration is leaded by the extension of a unique front lamellipodium. A thin extension is also visible at the uropod. The APC-silenced T cell migration pattern is rather characterized by the processive extension and retraction of membrane protrusions resulting in a not directional locomotion. This video is representative of 3 independent experiments. Time stamp is in min:sec. 20x/0.4 objective. Scale bar 10µm.
